# Association of Bassoon (BSN) Gene Mutations with Gait and Motor Impairments in Parkinson’s Disease

**DOI:** 10.1101/2025.08.10.25333397

**Authors:** Prashanth Lingappa Kukkle, Ahamed P Kaladiyil, Thenral S Geetha, Ramesh Menon, Rukmini Mridula Kandadai, Vinay Goyal, Soaham Dilip Desai, Deepika Joshi, Hrishikesh Kumar, Pettarusup M Wadia, Adreesh Mukherjee, Niraj Kumar, Sahil Mehta, Sandeep Chargulla, Shaktivel Murugan, Heli S Shah, Vijayshankar Paramanandam, Mitesh Chandarana, Ravi Yadav, Rajinder Dhamija, Pramod Kumar Pal, Atanu Biswas, Ravi Gupta, Rupam Borgohain, Vedam Ramprasad, Parkinson Research Alliance of India (PRAI)

## Abstract

**Introduction:** Parkinson’s Disease (PD) features debilitating motor symptoms, particularly gait and balance impairments inadequately managed by current therapies. Bassoon (BSN), a presynaptic active-zone organizer, has been implicated in various neurological disorders. Here, we evaluate the impact of rare BSN mutations on motor symptoms in PD patients.

**Methods:** Our study included 110 PD patients carrying BSN mutations and 558 PD controls from a South Asian early-onset PD cohort (onset <50 years). Variants with mean allele frequency (MAF) <0.1% were classified as “rare” (n=44). Clinical motor features were compared between variant carriers and non-carriers. Computational tools (CADD, PolyPhen-2, I-Mutant2.0, ConSurf) predicted deleteriousness, while GeneMANIA and STRING elucidated Bassoon’s functional interactions.

**Results:** Patients carrying BSN variants exhibited significantly increased freezing of gait (FOG, p=0.026, Carmer’s V=0.118), shuffling gait (SG, p=0.041, Carmer’s V=0.111), and falls (p=0.028, Carmer’s V=0.117). Rare BSN mutations clustered in the Bassoon C-terminal region (aa 3500– 3800), threefold above expected frequency. Computational predictions identified seven likely pathogenic variants (P171L, A852T, P988A, R1015H, R2561H, R3400W, L3561P), with highest confidence for P171L (confirmed by AlphaMissense). Functional analyses implicated Bassoon in axonal transport, presynaptic proteostasis, and neurotransmitter release in dopaminergic/cholinergic neurons.

**Conclusion:** Our findings identify BSN mutations as a genetic risk factor for PD-related gait and balance dysfunction, highlighting Bassoon’s role in neurotransmission. The link with Progressive Supranuclear Palsy phenotypes suggests Bassoon dysfunction could represent a convergence point between synucleinopathies and tauopathies.

## Introduction

Parkinson’s disease (PD), the second most common neurodegenerative disorder, encompasses a broad spectrum of symptoms that extend well beyond its hallmark triad of tremor, bradykinesia, and rigidity. Recent evidence demonstrates that PD pathophysiology is not confined to dopaminergic pathways; rather, multiple circuits and molecular mechanisms are implicated.^1–3^ This complexity is evident in the diverse clinical subtypes of PD, often classified by predominant motor or nonmotor features and distinct treatment responses.

Among the motor manifestations, gait and balance disturbances—especially freezing of gait and postural instability—are particularly debilitating. Although they typically emerge later in the disease course, some individuals experience these issues early and display poor responsiveness to levodopa, suggesting additional or alternative pathophysiological pathways.^4,5^ A notable parallel exists in Progressive Supranuclear Palsy (PSP), a tauopathy characterized by prominent early gait and balance deficits. Despite differing neuropathology (tau in PSP vs synuclein in PD), these two disorders frequently share clinical features.^6^

Intriguingly, the *BSN* gene, encoding the presynaptic protein Bassoon, has been linked to PSP-like syndromes in case studies presenting severe gait freezing and instability.^7^ Rare *BSN* variants have also been reported in epilepsy, multiple system atrophy, bipolar disorder, and schizophrenia.^8–10^ Whether these mutations represent incidental findings or a genuine pathogenic driver in parkinsonian syndromes remains uncertain. Our previous review article gives a comprehensive view of the current body of literature on Bassoon’s associations with various neurological diseases.^11^

In our recent gene-screening work on young-onset PD, *BSN* emerged as one of the top genes enriched for rare, predicted-deleterious variants, pointing to a role in disease susceptibility (p ≈ 2.5×10^–5).^12^ The identification of genetic associations of *BSN* gene with PD and PSP, diseases with overlapping phenotypes, led us to search for Bassoon’s role in parkinsonian pathology. Here, we identified a higher risk of gait dysfunction and falls in PD patients carrying *BSN* variants. This is the first report of a potential link between Bassoon and motor functions.

### Methodology

#### Patient Cohort

The study cohort consisted of early-onset Parkinson’s disease patients enrolled in a multi-center genetic study across India (GOPI-YOPD). In total, *n* = 668 PD patients with an age at onset ≤50 years (covering juvenile, young-onset, and early-onset PD) were recruited from movement disorder clinics nationwide between 2016 and 2020. This sample size is appropriate for establishing correlations between genotype and phenotype. Detailed methodology and key demographic features are provided in these publications.^12–14^ All participants provided informed consent, and the study was approved by the appropriate institutional ethics committees.

#### Genetic Screening and Variant Classification

Genomic DNA was extracted from blood samples of each patient. Whole-exome sequencing (WES) was performed on the cohort (668 PD cases). The sequencing and variant calling pipeline have been described previously.^12,14^ For the present analysis, we extracted all variants in the coding region of the *BSN* gene. Detected *BSN* variants included only missense mutations. Patients were grouped as BSN positive if they harbored any *BSN* mutation and BSN negative if they carried no *BSN* variants. We used the South Asian Genomic Research database (SARGAM) as a reference for allele frequencies. Variants with MAF <0.1% were categorized as “rare”. Among rare variants, we further subclassified them by rarity: “slightly rare” (MAF 0.05–0.1%), “moderately rare” (MAF 0.01–0.01%), and “Ultra rare” (MAF <0.01%).

#### Clinical Evaluation of Motor Impairment

Motor symptoms at presentation were recorded, including the presence or absence of tremor, rigidity, bradykinesia, dystonia, Asymmetric onset, cognitive impairment, and gait/postural impairment. We noted whether patients had a history of freezing of gait (FOG), shuffling gait (SG), and falls. These gait-related features were assessed through patient interviews and neurologic examination: FOG was identified by patient description of “feet glued to the floor” episodes, SG by observation of short stride length/festination, and falls by history of balance-related falls in the past. These observations were recorded as binary results.

#### Statistical Analysis

We assessed the association between *BSN* mutation status and motor impairments using the chi-square test (χ²). A p-value <0.05 was considered statistically significant. We also calculated Cramer’s V to estimate the effect size of the association (where V=0 indicates no association and V=1 indicates a perfect association). Additionally, we computed odds ratios (OR) with 95% Confidence Intervals (CI) to quantify the direction and strength of the associations.

All statistical analyses were conducted using SPSS software, and graphs were generated in Python. The use of proportional imputation on missing variables in different categories resulted in very similar of p and Cramer’s V. Dot plots plotting p values and Cramer’s V were generated to identify interactions between variables. We also constructed forest plots to visualize the ORs and 95% Confidence intervals for k ey associations (e.g. *BSN* carriers’ odds of FOG, SG, and falls, compared to non-carriers). In addition to group comparisons, we examined whether the frequency of rare *BSN* variants clustering in certain protein regions exceeded expected values (using binomial probability calculations).

#### Prediction of Functional effects of mutations

We used the following computational tools for prediction of pathogenicity of variants: CADD (Combined Annotation Dependent Depletion) to obtain a deleteriousness score for each gene variant, PolyPhen-2 (both HumVar and HumDiv models) to classify amino acid substitutions as benign or possibly/probably damaging, I-Mutant2.0 to calculate changes in ΔG between mutant and WT protein structure (ΔΔG), ConSurf to identify extent of evolutionary conservation, and AlphaMissense (an AlphaFold2-based deep learning model) to predict the functional effect of missense changes.^15–19^ Variants were considered *pathogenic* if they scored above established thresholds (CADD ≥20), were labeled as “possibly/probably damaging” by PolyPhen-2, and classified as “pathogenic” by AlphaMissense.

#### Visualization of Molecular Interactions

GeneMANIA tool was used to predict the biological functions of a gene based on genetic interactions, pathways, co-expression, co-localization, and similar protein domains.^20^ The Search Tool for the Retrieval of Interacting Genes (STRING) software enabled the visualization of protein-protein interaction networks as correlations between proteins. We also identified the enrichment of Bassoon in different Reactome pathways using STRING.^21^

## Results

### Clinical and Genetic Characteristics of *BSN* Mutation Carriers vs Non-Carriers

Out of the 668 PD patients in our cohort, 110 patients (16.3%) carried at least one mutation in the coding region of *BSN* gene (*BSN* positive), while the remaining 558 patients had no *BSN* variants (BSN negative). The mean age at onset of the patients was ∼39 years in both *BSN* positive and *BSN* negative patients, reflecting the early-onset nature of the cohort. One notable difference was the sex distribution: the *BSN* mutation carrier group had a lower male-to-female ratio (1.7:1) compared to the non-carrier group (2.3:1). The patients were followed up for an average of 95 months, with rigorous documentation of clinical features during the disease progression. The frequencies of classic PD motor symptoms (tremor, rigidity, bradykinesia, Asymmetric Onset) at presentation were high in both groups and did not differ significantly, suggesting that the *BSN* positive and *BSN* negative patients were broadly comparable in general PD features (Table 1) (Figure 1A).

**Figure 1:**
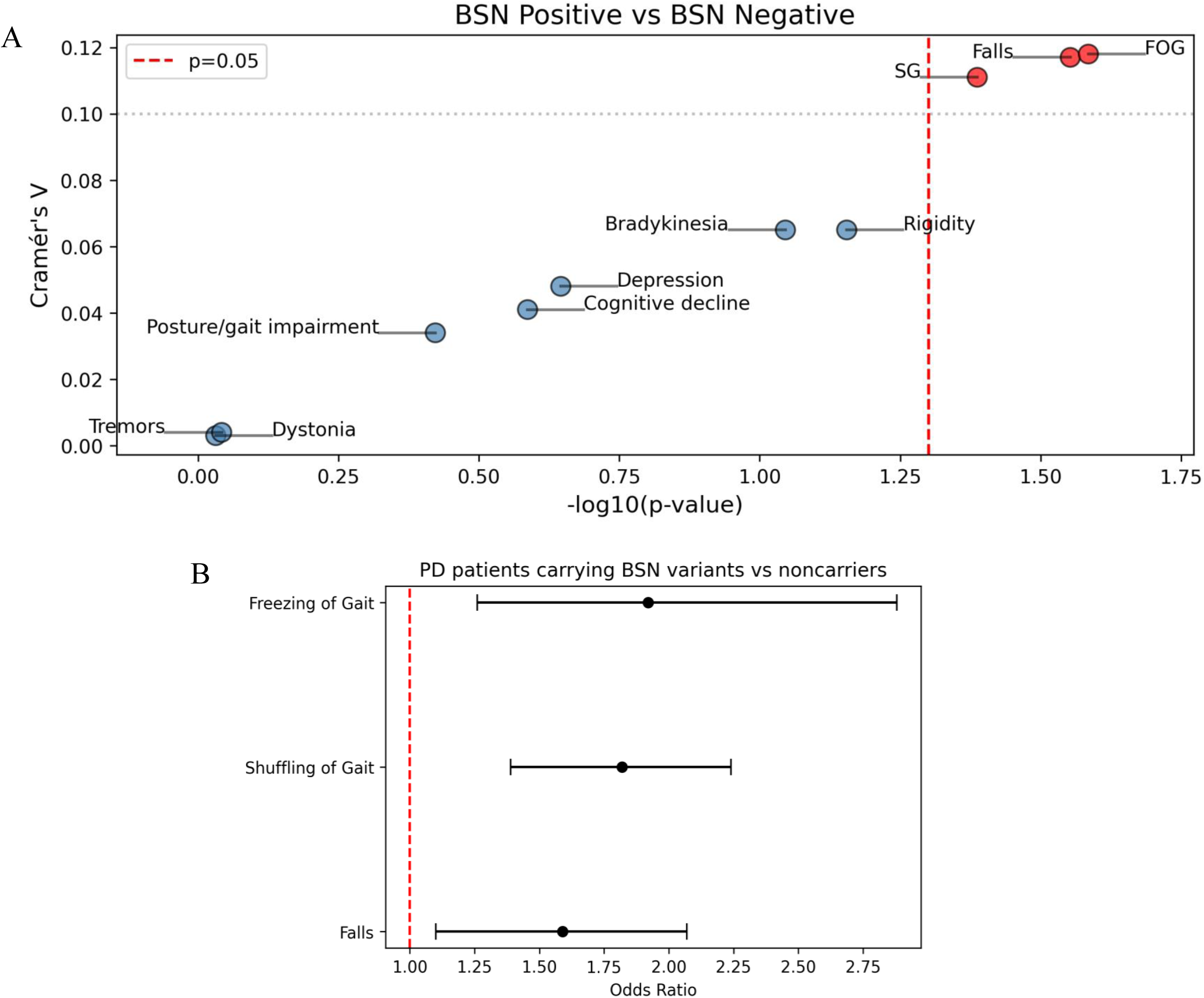
Phenotype analysis identifies freezing of gait, shuffling gait, and falls as correlated with BSN mutations in PD patients. A-Dot plot of p value and Cramer’s V indicating interactions between clinical phenotypes and mutations in *BSN* gene. Red dots indicate phenotypes with p<0.05 and Cramer’s V>0.1 indicating significant moderate association. B-Forest plot displaying odds ratio of BSN positive over BSN negative patients for developing FOG, SG, and Falls with a 95% confidence interval.

**Table 1:** Demographics and clinical reports of Parkinson’s Disease patients harboring mutations in *BSN* gene.

|  | *BSN* Positive | *BSN* Negative |
| --- | --- | --- |
| Demographic Features |  |  |
| No. of Subjects | 110 | 558 |
| Male to female ratio | 2 | 2.3 |
| Mean age at onset, years<br>(Range) | 39.1± 8.9<br>(14-70) | 39.4±8.7<br>(11-67) |
| Geographical distribution |  |  |
| North India | 32.4% | 31.8% |
| South India | 37% | 36.8% |
| East India | 16.6% | 22.1% |
| West India | 13.8% | 9.17% |
| Family history of PD (%) | 19% | 21.2% |
| **<u>Clinical Features at Presentation</u>** |  |  |
| Tremors (%) | 76.2% | 75.5% |
| Rigidity (%) | 68.6% | 79.4% |
| Bradykinesia (%) | 83.8% | 90.7% |
| Postural/Gait impairment (%) | 57.3% | 53.7% |
| Dystonia (%) | 19% | 21.1% |
| Asymmetric onset (%) | 96.2% | 95.6% |
| Memory/cognitive deterioration | 18.5% | 23.7% |
| **<u>Polygenic Risk Score (PRS)</u>** |  |  |
| Low risk patients (number of patients) | 18 | 59 |
| High risk patients (number of patients) | 27 | 182 |

Crucially, however, a higher proportion of *BSN* positive patients exhibited the PIGD phenotype (postural/gait impairment) (57.1% in *BSN* positive vs 53.7% in *BSN* negative) (Table 1). More specifically, we found that freezing of gait, shuffling gait, and falls were all significantly more common in *BSN* positive PD patients than in those without *BSN* mutations (Figure 1A). FOG was reported to be significantly higher in *BSN* carriers (*p = 0.026, Cramer’s V = 0.118*) and a moderate association was observed. SG was also more frequent in *BSN* positive patients (*p = 0.041, Cramer’s V = 0.111*), and a history of falls was more common in *BSN* positive patients (*p = 0.028, Cramer’s V = 0.117*). Although the effect sizes (*Cramer’s V ∼0.11–0.12*) were modest, the consistent pattern across these features suggests that carriers of *BSN* mutations showed a discernible increase in gait and balance impairment relative to non-carriers. To further illustrate the magnitude of these associations, we calculated the odds ratios for *BSN* positive patients developing each symptom. The odds of experiencing FOG were 1.92-fold higher for *BSN* mutant patients compared to *BSN* negative patients. Similarly, *BSN* mutation carriers had an *OR = 1.82* for developing SG, and *OR = 1.59* for having recurrent falls. All OR 95% confidence intervals excluded 1, confirming a significant positive correlation between *BSN* variants and these motor deficits (Figure 1B). A maximum of 16 missing values was found in each clinical variable (4 in *BSN* positive and 12 in *BSN* negative). Taken together, these results establish that PD patients carrying *BSN* mutations are more prone to FOG, SG, and falls than those without *BSN* mutations, implicating the role of *BSN* gene in these motor phenotypes.

### Rare *BSN* Variants Contribute to Impairments in Posture Functions

To refine the analysis, we next examined whether the association with motor deficits was driven primarily by rare, potentially pathogenic *BSN* variants as opposed to common benign variants. Among the 110 *BSN* positive patients, 44 patients carried rare *BSN* mutations (MAF < 0.1%), while the remaining 66 patients had only common *BSN* variants (MAF ≥ 0.1%). Table 2 summarizes the demographic and clinical features of these subgroups which remained largely similar. PD Patients with rare *BSN* variants tended to be predominantly male (male: female ratio of 3:1) compared to those with common variants (1.3:1). Importantly, the rare-variant carriers showed a higher frequency of gait defects. 61.3% of rare-*BSN* carriers had postural instability/gait difficulty at presentation, in comparison to 54.6% of the common-variant group (table 2). We compared the rare-*BSN* group against the *BSN* negative group to test for associations in this enriched subset. Among the clinical phenotypes described earlier, only FOG and SG were found to have significant associations with the presence of rare *BSN* variants (Figure 2A). FOG was present at a significantly greater frequency in the rare-*BSN* group (*p = 0.005, Cramer’s V = 0.115*), giving an odds ratio of 2.59 vs *BSN* negative patients. SG was also markedly associated with rare *BSN* variants (*p = 0.018, Craver’s V = 0.097*), with an OR of 2.31 for rare-variant carriers relative to *BSN* negative (Figure 2B). These ORs for FOG and SG in the rare variant subset were higher than those observed for *BSN* mutations overall, indicating that the rare, low-frequency *BSN* mutations confer a greater risk for gait dysfunction when common variants were excluded. In contrast, falls did not show a significant association in the rare-variant subset (*p = 0.46*), although a larger percentage of rare-variant carriers with history of falls was observed compared to *BSN* negative (24.3% vs 18.4%). This could suggest that the link between *BSN* and falls observed in the combined analysis might be driven by a broader set of variants or indirect factors. It is also possible that with the smaller sample size of rare variant carriers (n=44), there was limited statistical power to detect a difference in falls, or that common *BSN* variants (or intermediate-frequency variants) have some contributory effect on fall risk. Nevertheless, the rare variant analysis clearly implicates rare *BSN* mutations as the key drivers of the observed motor associations.

**Figure 2:**
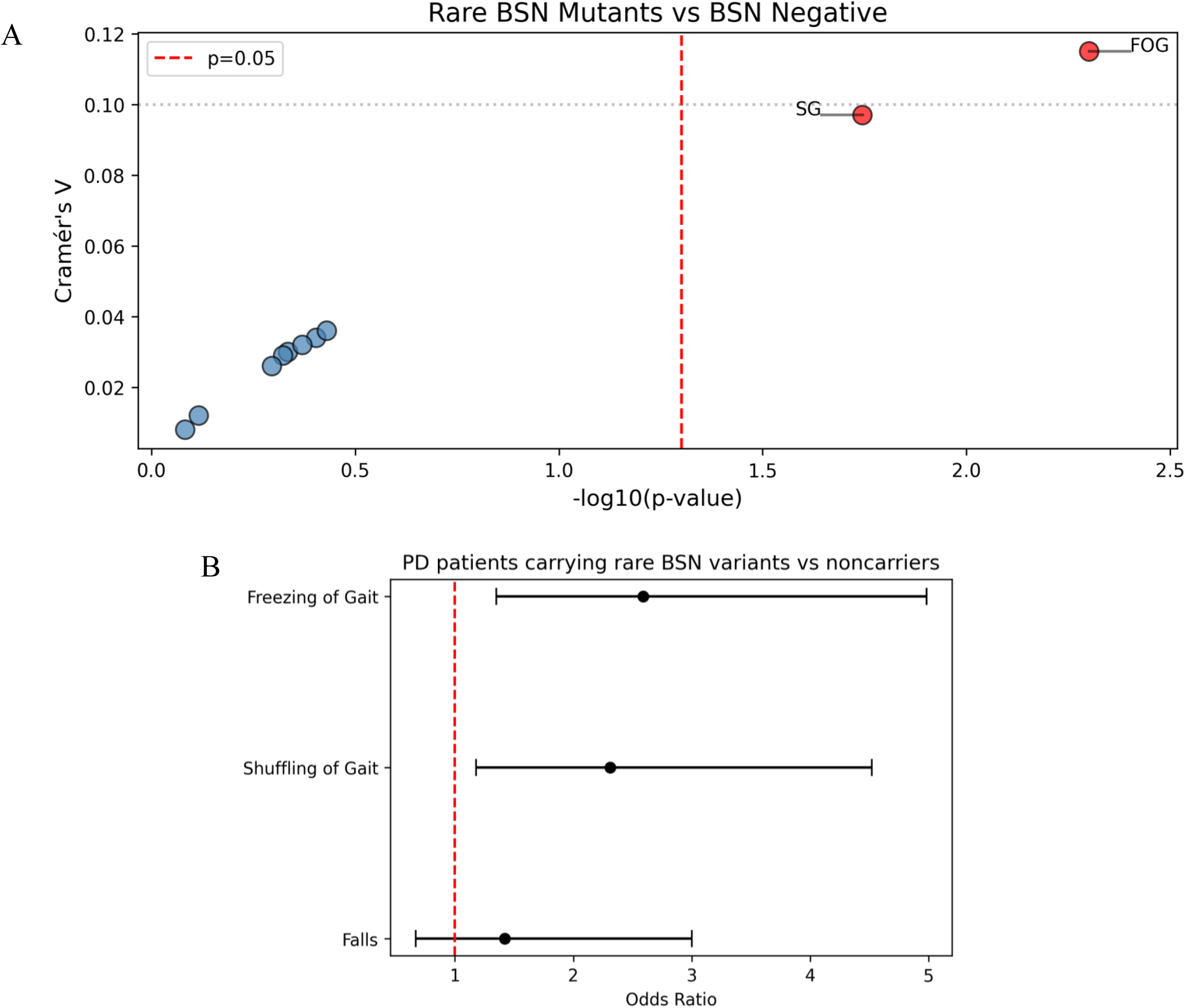
Rare BSN mutation carriers display stronger association with gait defects. A-Dot plot of p value and Cramer’s V indicating interactions between clinical phenotypes and mutations in *BSN* gene. Red dots indicate phenotypes with p<0.05 and Cramer’s V>0.1 indicating significant moderate association. B-Forest plot displaying odds ratio of rare BSN variant carrying patients for developing FOG, SG, and Falls with a 95 % confidence interval.

**Table 2:** Demographics and clinical reports of Parkinson’s Disease patients harboring rare and common mutations in the *BSN* gene. ^#^ “Rare BSN Mutants”- PD patients carrying variants in BSN gene of minor allele frequency <0.1% across the databases “ExAC, 1000Genome, (v3.1), gnomAD(v2), gnomAD(v2), Topmed, Genome Asia, SAS-ATLAS”

|  |  |  |
| --- | --- | --- |
|  | **Rare *BSN* mutants<sup>#</sup>** | **Common *BSN* mutants** |
| **Demographic Features** |  |  |
| No. of Subjects | 44 | 66 |
| Male to female ratio | 3 | 1.3 |
| Mean age at onset, years<br>(Range) | 38.7 ± 1.9<br>(14-70) | 39.0 ± 1.2<br>(11-67) |
| Age at onset<br>11 - 20 years<br>21 - 30 years<br>31 - 40 years<br>41 - 50 years<br>>50 years | 04 (3.8%)<br>10 (9.5%)<br>41 (39%)<br>46 (43.8%)<br>04 (3.8%) | 17 (3.0%)<br>57 (10.1%)<br>210 (37.5%)<br>252 (45.0%)<br>23 (4.1%) |
| Geographical distribution<br>North India<br>South India<br>East India<br>West India | 34%<br>38.6%<br>20.4%<br>6.8% | 30.7%<br>35.3%<br>13.8%<br>18.4% |
| Family history of PD (%) | 16.2% | 19.6% |
| **<u>Clinical Features at Presentation</u>** |  |  |
| Tremors (%) | 84% | 80.3% |
| Rigidity (%) | 75% | 67.6% |
| Bradykinesia (%) | 86.3% | 83% |
| Postural/Gait impairment (%) | 61.3% | 54.6% |
| Dystonia (%) | 26.8% | 20.6% |
| Asymmetric onset (%) | 95.4% | 96.9% |
| Memory/cognitive deterioration | 20.9% | 16.9% |
| **<u>Polygenic Risk Score (PRS)</u>** |  |  |
| Low risk (number of patients) | 6 | 12 |
| High risk (number of patients) | 13 | 16 |

Consistently, when we examined the subgroup of 66 patients with common *BSN* variants (MAF>0.1%), we found no significant difference in FOG, SG, or falls compared to *BSN* negative patients, implying that common *BSN* polymorphisms did not impact gait or balance in PD (Table 3). This reinforces the interpretation that *BSN*’s influence on the PD motor phenotype is likely mediated by rare potentially pathogenic mutations rather than common or tolerated genetic variations.

**Table 3:** Percentage of PD patients with different rarity of *BSN* mutations exhibiting motor defects. 44 rare variants were further subcategorized based on minor allele frequencies as a) Slightly rare (0.1% to 0.05%) b) Moderately rare (0.05% to 0.01%) c) Ultra rare <0.01%. The percentage of patients within these subsets exhibiting FOG, SG, and Falls were compared.

| Motor Features | Ultra rare *BSN* variant (n = 15) | Moderately rare *BSN* variants (n = 13) | Slightly rare *BSN* variant (n = 16) | Common *BSN* mutations (n = 66) | *BSN* negative (n = 558) |
| --- | --- | --- | --- | --- | --- |
| Freezing of gait (%) | 53.8% | 61.5% | 57.1% | 45.4% | 34.2% |
| Shuffling gait (%) | 66.6% | 69.2% | 75% | 61.5% | 50.7% |
| Falls (%) | 30.7% | 25% | 18.7% | 26.9% | 18.4% |

We also explored whether increasingly stringent rarity of *BSN* variants correlated with more severe phenotypes. Although sample sizes became small when subdividing the 44 rare mutation carriers, we observed a trend: the percentage of patients who had frequent experiences of falls appeared to increase as *BSN* variant frequency decreased (Table 3). For FOG and SG, the percentages were high across all rare subgroups with no clear trend following rarity. This pattern raises the possibility that extremely rare *BSN* mutations – likely to be the most deleterious – might contribute not only to gait freezing but also to postural instability to a greater extent. This remains a tentative observation that would require a larger dataset to verify.

### In Silico Predicted Impact of *BSN* Mutations

Beyond the clinical correlations, we made an interesting observation in the distribution of rare *BSN* mutations along the Bassoon protein. On mapping the 44 rare *BSN* variants onto the Bassoon protein sequence, we noticed a cluster of mutations in the C-terminal region of Bassoon (Figure 3A). Specifically, 9 out of the 44 rare-variant carriers (20.4%) had mutations located between amino acid positions 3500 and 3800 of the Bassoon protein. This 300-amino acid residue segment corresponds to the C-terminus of Bassoon. We computed the probability of this clustering occurring by chance, given that the Bassoon protein is 3926 amino acids long, to be 7% (200/3926 ≈ 0.07). Yet in our data ∼20% of rare variants occurred in this C-terminal stretch, a frequency far exceeding random expectation (binomial probability *p 0.01*). Thus, the C-terminal region of Bassoon, known to mediate functions like tethering synaptic vesicles to the Active Zone (AZ), appears to be a hotspot for rare mutations in PD patients. A similar enrichment of epilepsy and neurodevelopmental disorders-associated variants in Bassoon’s C-terminus was reported by Ye et al.^8^ This implies the functional criticality of Bassoon’s C-terminal domain.

**Figure 3:**
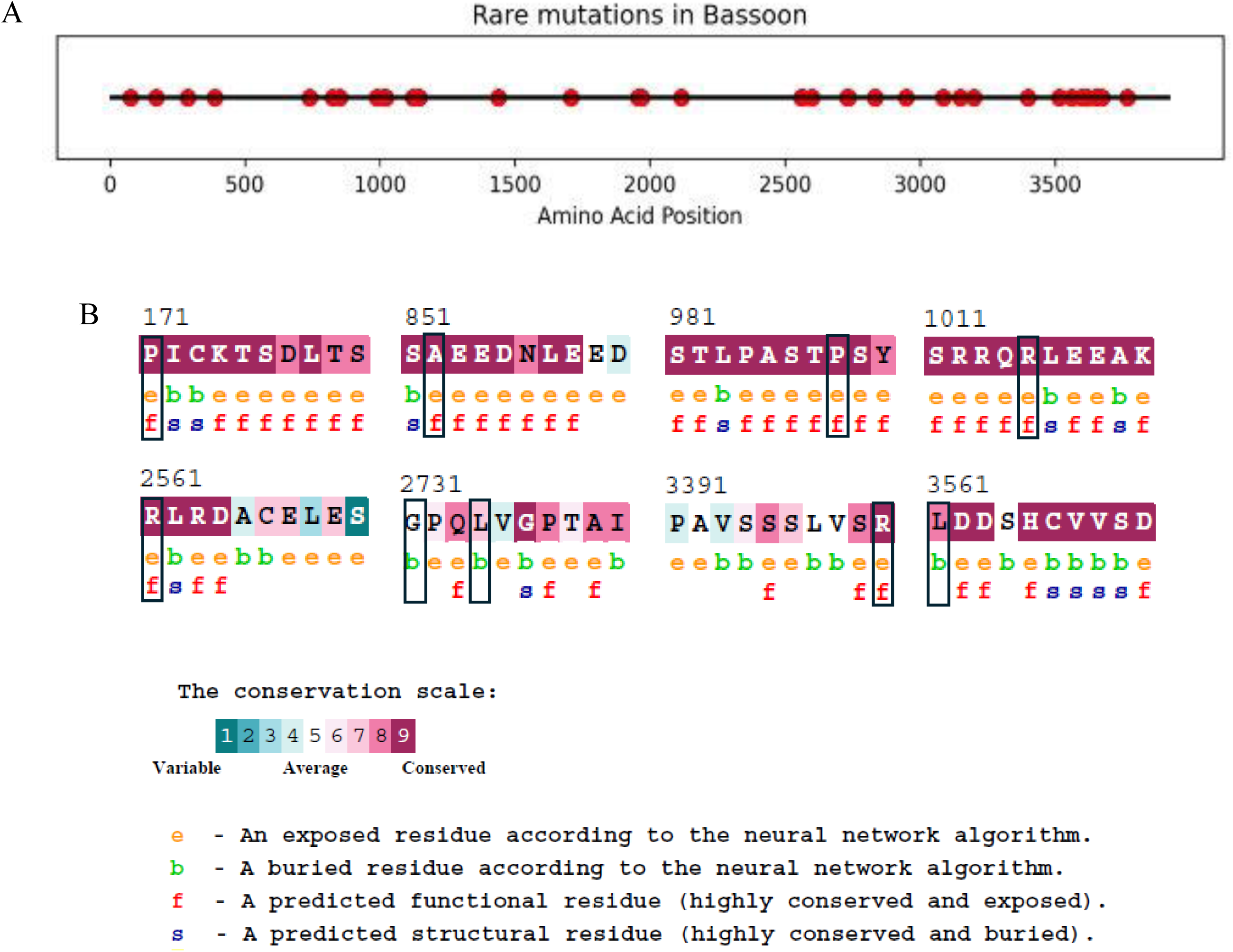
Potentially deleterious rare mutations in Bassoon. **A**-Line plot visualizing locations of rare mutations in the Bassoon protein. B-Amino acid residues P171, A852, P988, R1015, R2561, R3400, L3561 are scored 7 or 8 by ConSurf, indicating high evolutionary conservation, whereas residues G2731 and L2734 score 5 or 6, implying moderate conservation.

To gain insight into the *BSN* variants that might be functionally consequential, we performed predictive analyses of the rare mutations identified. Using multiple computational tools, we evaluated the potential structural and functional impact of each variant.

First, we used the CADD score, which integrates diverse genomic features to estimate variant deleteriousness. We found that 32 out of the 35 distinct *BSN* variants had CADD scores >20, a common threshold for pathogenicity. We then proceeded to use the HumVar model of the PolyPhen-2 tool which classified 24 out of the 35 mutations in the Bassoon protein as possibly or probably damaging. These 24 mutations were also predicted to be damaging by the HumDiv model of PolyPhen-2. Of these 24, 22 were also classified by CADD as potentially pathogenic (CADD > 20), showing a significant overlap in classification. It is notable that several of these PolyPhen2-flagged mutations lie in functionally important domains: for instance, R3515C and L3561P are in the Bassoon C-terminus near the mutation cluster.

Further, we used the I-Mutant2.0 software to calculate the ΔΔG values for the 22 mutations identified by CADD and PolyPhen2 as damaging. We found 9 mutations to have a highly negative ΔΔG value (less than -0.8); P171L, A852T, P988A, R1015H, R2561H, G2731R, L2734R, R3400W, and L3561P (Table 4). This enabled us to select for mutations that have a strong destabilizing effect on overall protein structure and stability.

**Table 4:**
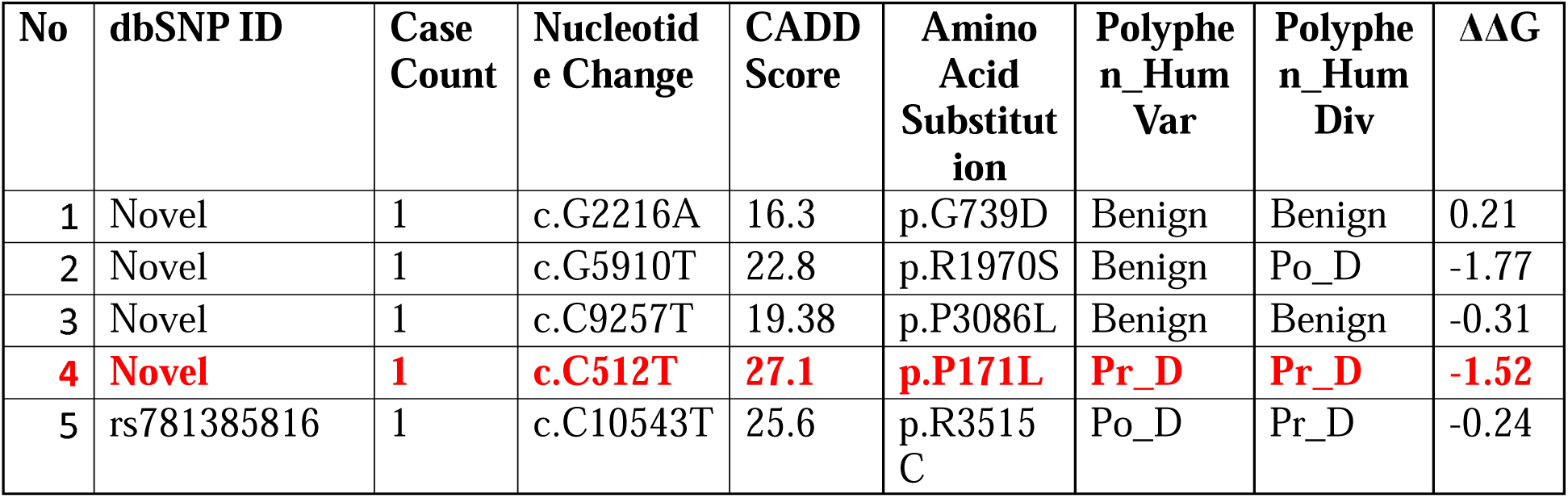
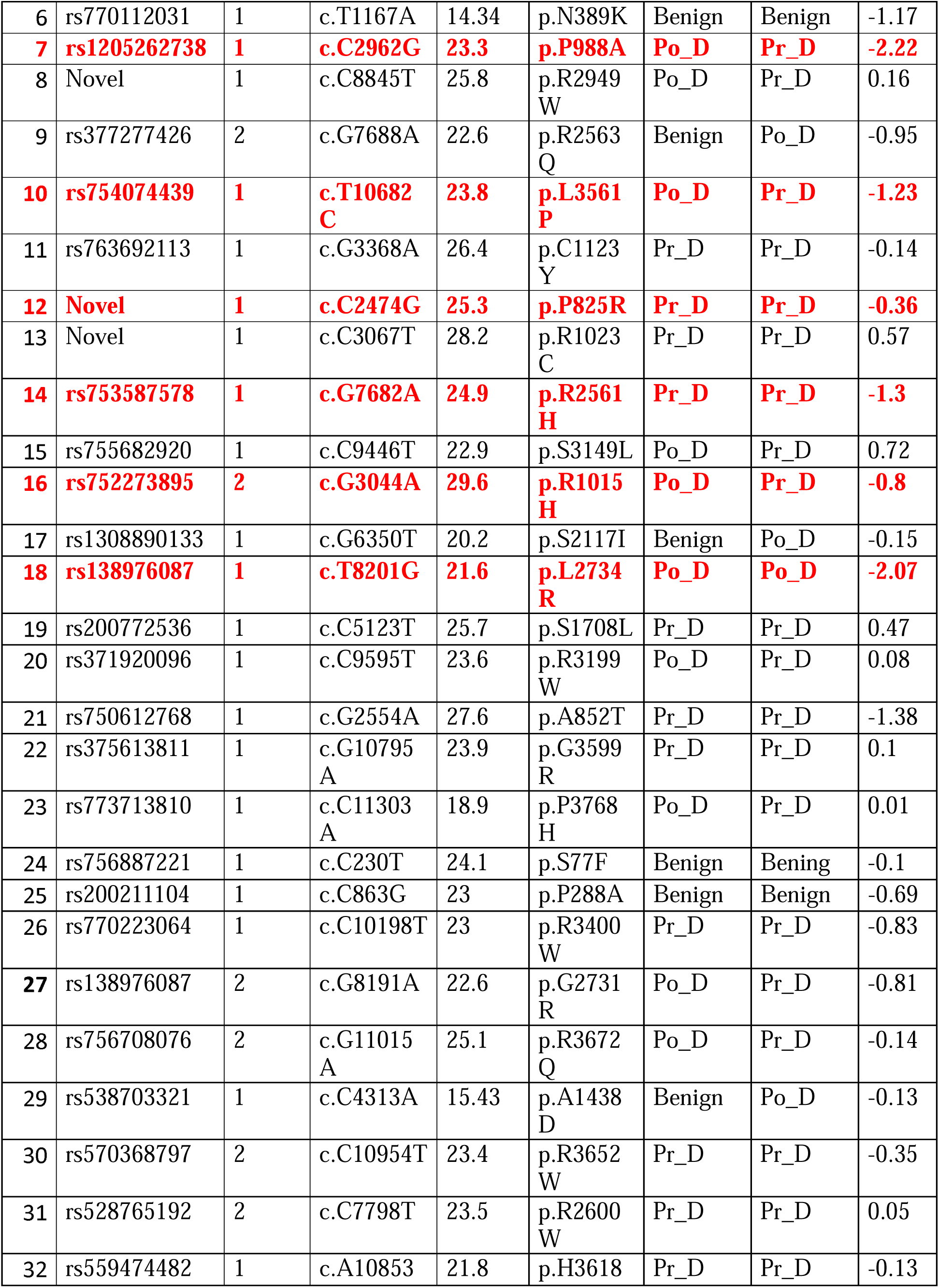
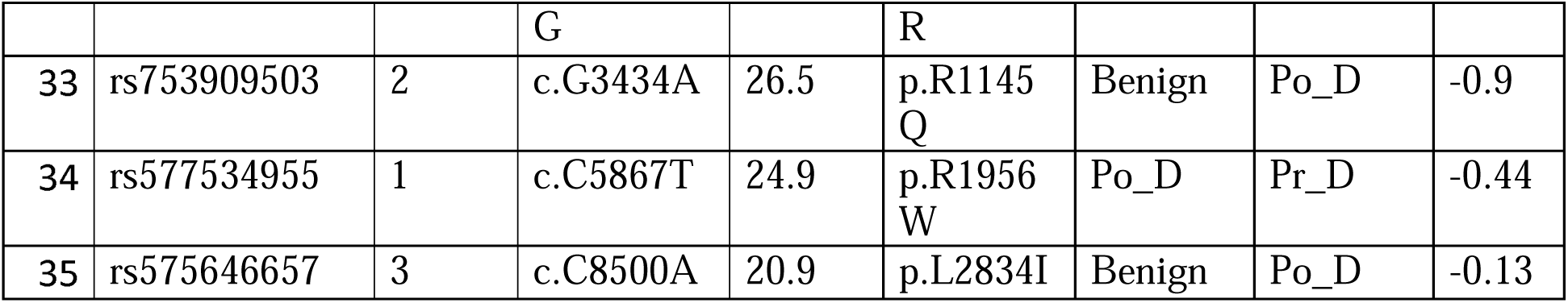
44 rare *BSN* mutations of MAF <0.1% observed in PD patients. Mutations are listed in increasing order of MAF. CADD scored variants from 0 to 30, higher score indicating increasing deleteriousness. PolyPhen2 classified mutations as benign (0.00-0.45), possibly damaging (0.45-0.95), probably damaging (0.95-1). I-Mutant2.0 scored mutations between –1 and 1 according to ΔΔG values, negative score indicating destabilizing effects and positive scores as stabilizing effects on protein structure. Mutations predicted to be likely damaging are highlighted in red. Abbreviations: Po_D: Possible damaging, Pr_D: Probably damaging

Using the ConSurf tool, we studied the evolutionary conservation of each of these 9 sites. Figure 3 shows that 7 out of the 9 sites are highly conserved, with a conservation scale value of 8 or 9, calculated based on phylogenetic relations between homologous sequences (P171, A852, P988, R1015, R2561, R3400, and L3561). The residues G2731 and L2734 were found to have average and slightly conserved scores respectively. Most of these residues were predicted to be exposed and functional (Figure 3). Interestingly, all 8 patients carrying the 7 potentially damaging mutations in conserved residues of Bassoon displayed impairments in posture or gait, further strengthening the role of Bassoon in posture and gait functions.

A new deep learning model built on AlphaFold, AlphaMissense, has been shown to outperform most previous models. This software predicted only one mutation, P171L, to be pathogenic. The P171 residue lies near the N-terminus of Bassoon in its Zinc-Finger domain, a well-structured domain involved in protein-protein interactions.

Interestingly, this mutation was from the highly rare subgroup and among the ones identified with a CADD score >25 and classified by PolyPhen-2 HumVar and HumDiv as probably damaging, yielding a consensus across tools. While computational predictions have their limitations, the consensus between different tools increases confidence that these mutations are likely to alter Bassoon’s structure and function.

### Functional Annotation of Bassoon

To elucidate the functional interaction network of the *BSN* gene, we used the GeneMANIA tool to predict the interactions with related genes. We identified strongest interactions of *BSN* with genes that are essential for synaptic vesicle exocytosis and recycling such as PCLO, ERC2, UNC13A and RIMS1. We also found an association with genes involved in synaptic signaling (RGS7, CAMKK1, and PPP6RA) (Figure 4A).

**Figure 4:**
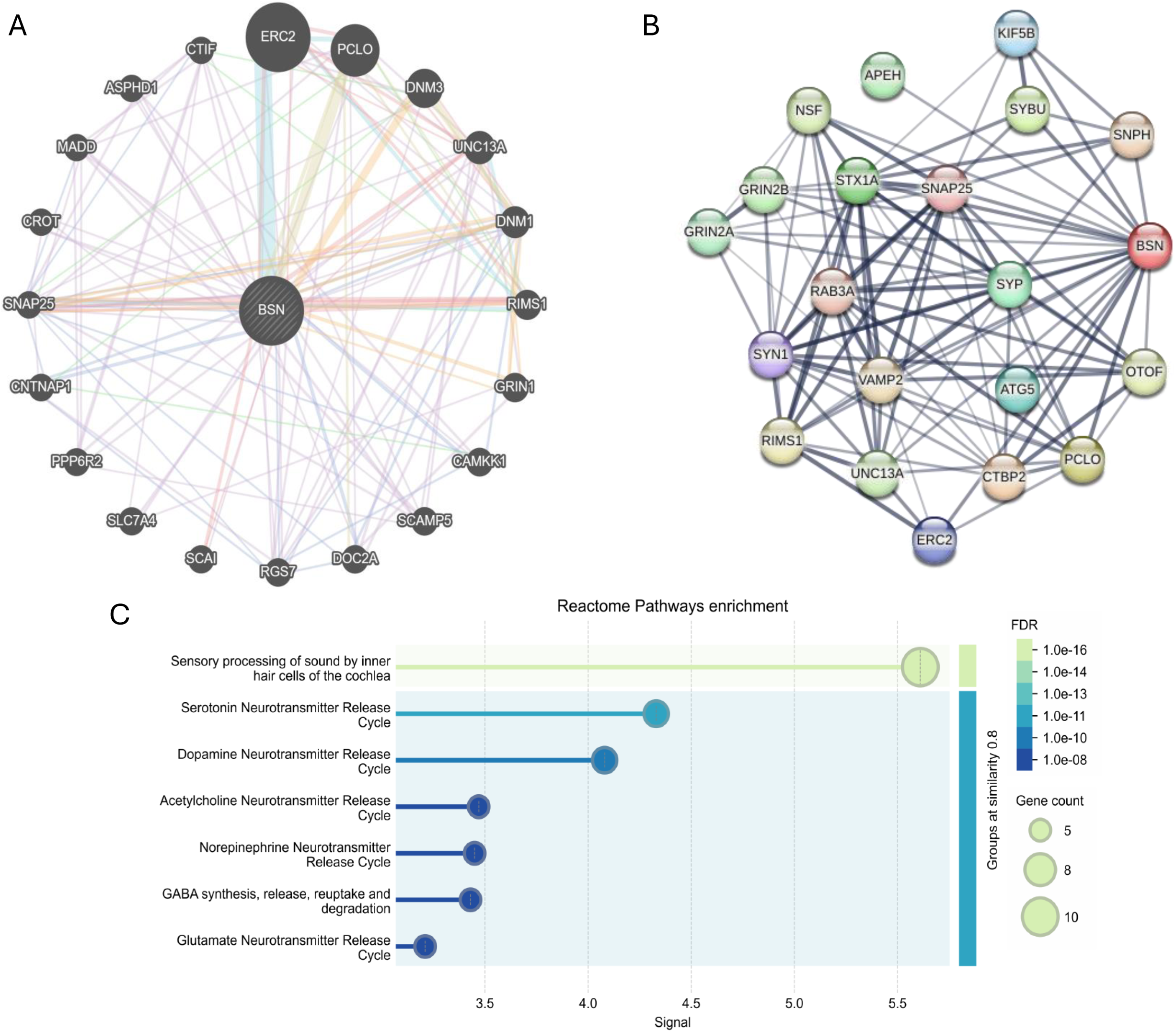
Gene-gene and protein-protein interaction networks of Bassoon. A-Gene interaction network of *BSN* as predicted by GeneMANIA tool. GeneMANIA web tool was utilized to generate the image. B-Protein interaction network of Bassoon, predicted by STRING tool. Image generated using STRING web interface. C-Enrichment of Bassoon in reactome pathways. Plot generated using STRING web interface.

A network of Bassoon’s protein-protein interactions was built using the STRING database to identify functional relationships with other proteins. Notably, Bassoon exhibited strong interactions with proteins involved in anterograde axonal transport, including KIF5B, NSF, and SYBU, suggesting a potential role in regulating the transport of synaptic components. Additionally, Bassoon was found to interact with proteins critical for SNARE complex formation, such as STX1A, SNAP25, SNPH, and VAMP2, highlighting its involvement in synaptic vesicle docking and fusion. Further analysis identified interactions with proteins essential for synaptic vesicle exocytosis, including OTOF, RIMS1, and UNC13A, underscoring Bassoon’s role in modulating neurotransmitter release. Bassoon’s function in synaptic vesicle reloading was also supported by its interactions with PCLO, RAB3A, and SYP, which are known to participate in vesicle recycling and replenishment. Interestingly, Bassoon’s interaction network extended to proteins involved in maintaining proteostasis, such as ATG5 and CTBP2, suggesting a potential role in protein quality control and degradation pathways within the synapse. These findings collectively highlight the multifaceted roles of Bassoon in synaptic function and regulation (Figure 4B).

Reactome pathway classification of Bassoon’s functions and roles pointed to an overall involvement of Bassoon in various neurotransmitter release, including dopamine, Acetylcholine, GABA, Serotonin, and Norepinephrine. This further strengthens Bassoon’s essential role in synaptic activity with a potential link to motor functioning. (Figure 4C).

In summary, our results demonstrate an association between *BSN* gene mutations and motor impairments in PD, particularly freezing of gait, shuffling gait, and increased falls. This association is driven mainly by rare *BSN* variants, some of which cluster in a critical C-terminal region of the Bassoon protein and are predicted to be damaging. We also identified potentially pathogenic missense mutations in Bassoon, linked to the development of the aforementioned phenotypes: P171L, A852T, P988A, R1015H, R2561H, R3400W, and L3561P, with a higher confidence in P171L. We also predict a significant role for Bassoon in neurotransmitter release in dopaminergic and cholinergic neurons, potentially impacting motor functioning.

## Discussion

We investigated the role of BSN gene variants in PD, identifying a significant association with gait and balance difficulties, particularly freezing of gait, shuffling gait, and falls. To our knowledge, this is the first clinical association linking BSN mutations to specific PD motor symptoms. Our findings suggest that Bassoon, traditionally unlinked to PD pathology, may act as a risk factor enhancing susceptibility to gait-related symptoms that are less dopamine-dependent.

### Mechanistic considerations

The link between *BSN* mutations and gait/motor impairment in PD can be considered from a neurophysiological perspective. Bassoon is a critical organizer of the presynaptic AZ, and it ensures efficient neurotransmitter release during high-frequency firing by tethering synaptic vesicles to the release sites. A plausible hypothesis is that Bassoon dysfunction leads to impaired synaptic transmission in motor control circuits, thereby manifesting as gait ignition failure (FOG) and poor postural control. One key circuit in PD **FOG** is the cortico-basal ganglia-brainstem loop involving the pedunculopontine nucleus (PPN), which is a cholinergic center regulating gait and posture. There is some evidence that Bassoon is present not only at glutamatergic and GABAergic synapses, but also at cholinergic synapses [22]. If Bassoon function in cholinergic PPN or related brainstem circuits are compromised, it could result in the gait deficits we observe. In fact, Bassoon mutant models provide insight into its importance: mice with a P3882A mutation in Bassoon have defects in locomotion, starting from 3 months of age [23]. In addition, Bassoon expressing flies also seemed to have motor deficits that reduced its climbing ability [24]. The absence of Bassoon has profound synaptic transmission defects, including an inability to sustain neurotransmitter release **on** repetitive stimulation, leading to an exaggerated presynaptic short-term depression. In inner hair cell synapses of the auditory system, Bassoon knockout led to rapid synaptic fatigue under high-frequency stimulation [25].

Transposing this idea to locomotor networks, Bassoon insufficiency might cause crucial synapses to depress or fail under continuous use, precipitating freezing episodes when a steady stream of neural signals is required to maintain walking. Additionally, Bassoon knockout mice exhibit neurological phenotypes like increased susceptibility to seizures and pre-weaning lethality, emphasizing that Bassoon is indispensable for normal network activity [8]. It stands to reason that heterozygous disruptive variants in *BSN* could reduce synaptic efficiency, affecting behaviors like gait.

Another angle is Bassoon’s role in neuronal proteostasis and protein aggregation. Bassoon has an unusually large structure and interacts with the ubiquitin-proteasome system components, implying its role in protein turnover [26,27]. If *BSN* mutations hamper this function, they might create a permissive environment for toxic protein accumulation, as seen in its role in tau aggregation [28]. Remarkably, Bassoon shares intriguing parallels with α-synuclein, the hallmark protein of PD. Like Bassoon, α-synuclein is involved in presynaptic vesicle dynamics and has been shown to modulate recovery from synaptic depression [29]. Certain mutations in the α-synuclein gene (*SNCA*) cause familial parkinsonism where patients present with early gait impairment and postural instability. For instance, the A53T α-synuclein mutation can lead to a syndrome with severe bradykinesia and balance problems, [30] and other *SNCA* variants have been linked to gait disorder in parkinsonian syndromes [31]. Both α-synuclein and Bassoon tend to aggregate abnormally: α-synuclein aggregates to form Lewy bodies in PD, and Bassoon has been observed to form presynaptic protein aggregates in certain conditions. Bassoon’s role in tau aggregation ties it to PSP. In the PSP-like disorder associated with a *BSN* mutation, [7] postmortem analysis indeed showed tau deposition in the brain, presumably triggered by the Bassoon mutation. Therefore, one can speculate that in PD, *BSN* mutations might also promote subtle increase in tau/α-synuclein aggregation and pathology that preferentially affects gait-related brain regions.

During our analysis, we find that rare *BSN* variants have stronger effects than common ones, likely due to their greater potential to disrupt protein structure. Since common variants are typically benign, it was mostly patients with rare function-perturbing *BSN* variants that exhibited significant gait impairment. In our cohort, a substantial fraction of the rare variants clustered in the Bassoon C-terminus, which is crucial for tethering synaptic vesicles to the active zone matrix. We hypothesize that mutations in Bassoon’s C-terminal region may weaken its ability to interact with AZ proteins such as Munc13 and RIM1, leading to inefficient synaptic transmission during continuous movement and thereby contributing to gait freezing. This is supported by the observation that most patients with C-terminal *BSN* mutations had severe PIGD symptoms. The clustering of disease-associated mutations in that region further points to its importance. However, caution is warranted – the human Bassoon protein is large and not well understood structurally, so pinpointing a mutation hotspot requires more data and possibly structural modeling to identify protein interaction sites.

To identify the mutations that could have drastic effects on Bassoon’s structure and function, we performed a systematic and detailed bioinformatic analysis. Using a range of predictive tools, we singled out the P171L mutation as likely pathogenic with high confidence. Although 6 other mutations were predicted to have deleterious effects by CADD, Polyphen, and I-Mutant2.0, the AlphaMissense tool picked out only P171L as likely pathogenic in its prediction. This increases the confidence for the P171L mutation. The use of SNPs&GO, a standard tool for predicting disease causing mutations, classified all 35 mutations as benign and was therefore not used in narrowing down the list of mutations. A similar tool, Panther, was not used for analysis here as it left a large number of mutations as unclassified.

The P171 residue lies in one of Bassoon’s few well-structured domains, the Zinc-finger domain. A proline-to-leucine change at this site could alter local structure and folding, enabled by proline’s rigid structure and hydrophobic interactions. This could drastically impact protein-protein interactions enabled by the ZnF domain. Although ZnF mediated protein interactions are not well studied for Bassoon, ZnFs of Piccolo, a structurally and functionally similar protein to Bassoon, were found to interact with synaptic vesicle associated proteins at the presynapse [32]. This leads us to believe that the ZnF of Bassoon, where the N-terminus is shown to interact with the synaptic vesicle, is likely involved in synaptic vesicle anchoring. The other 6 likely pathogenic mutations in Bassoon were scattered across the protein, rendering us unable to draw further conclusions on functional implications.

The Functional annotation of the Bassoon protein emphasizes a major role in presynaptic neurotransmission. Bassoon functions in anchoring synaptic vesicles to the AZ, regulating the priming and exocytosis of the vesicles. Besides this, Bassoon plays a role in protein localization to the presynapse through anterograde axonal transport and in maintaining proteostasis by regulating transcription and autophagy.

### Overlap with PSP and other disorders

Our results reinforce the bridge between PD and PSP. The *BSN* mutation carriers in PD exhibit gait and postural symptoms that are the hallmark early features of PSP. Conversely, *BSN* mutations in PSP-like syndrome induce tau pathology and severe gait impairment. It is tempting to speculate that Bassoon-related pathology might represent a point of convergence between α-synucleinopathy and tauopathy. Perhaps Bassoon dysfunction leads to synaptic protein mishandling that can trigger tau accumulation in susceptible individuals (leading to a PSP presentation) or exacerbate synuclein toxicity in others (contributing to PD). The fact that Bassoon can influence tau aggregation and that Bassoon, like α-synuclein, modulate presynaptic vesicle dynamics suggests a model where Bassoon is a central maintenance factor for synaptic integrity [28]. A *BSN* mutation carrier might develop PSP if they also have genetic predisposition to tauopathy, whereas in a PD background, the same mutation might worsen gait symptoms without changing the primary disease course. This could reason why *BSN* is not a known monogenic cause of PD or PSP, but rather a risk factor that modulates disease severity or phenotype. To ensure accuracy in diagnosis, we conducted long term follow ups of the PD patients, ruling out possibilities of classical or evolving PSP.

### Limitations of the Study

While our findings are statistically significant, *BSN* mutations have modest effect sizes (Cramer’s V ∼0.1), implying they are one of many factors influencing gait impairment. Freezing of gait (FOG) and falls in PD are multifactorial, influenced by disease duration, medication, severity, and comorbidities. Although we controlled for some variables, unmeasured confounders such as disease severity exists that cannot be accounted for due to our small sample size of *BSN* positive patients. Our cohort is ethnically homogeneous (Indian/South Asian), requiring replication in diverse populations, as different rare *BSN* variants may exist across groups. Since our study used Whole Exome Sequencing, we do not have information about intronic variants as well as variants in other transcriptional regulation sites that could regulate the expression of the protein. Future studies should analyze larger PD datasets (e.g., International Parkinson’s Genetics Consortium) to confirm findings. Finally, FOG, SG, and falls were based on clinical assessments and patient recall, which may introduce misclassification. Future research should incorporate objective gait analysis or wearable sensors for more precise phenotyping. However, our clinical measures remain robust for group comparisons. The use of predictive tools for characterizing missense mutations in the Bassoon protein risk misinterpretation or overlooking of critical mutations. This limitation stems from their inability to reliably predict known and unknown protein-protein interactions, particularly for proteins like Bassoon whose structure remains unknown. This may be why the tools used do not pick up on the functional C terminal domain of *BSN* as a hotspot for mutations.

### Future Directions

Future research should explore the biochemical relationship between Bassoon, tau, and α-synuclein, as well as functional validation of mutations in cellular and animal models. Investigations into larger PD datasets or atypical parkinsonism (PSP) cohorts could further validate BSN’s role [7,33]. Determining if BSN carriers display increased tau pathology or α-synuclein aggregation could clarify underlying mechanisms.

## Conclusion

Our findings implicate BSN mutations in gait and postural dysfunction in PD, presenting parallels with PSP phenotypes. These observations expand PD’s genetic landscape, introducing Bassoon as a novel factor influencing PD motor phenotypes. Bassoon may represent a critical node linking presynaptic dysfunction and protein aggregation pathways. If validated, therapeutic strategies targeting Bassoon-related pathways could address gait impairments poorly managed by current dopamine-based therapies. Ultimately, integrating genetics with neurobiology may advance personalized interventions for PD, encouraging broader explorations across synucleinopathies and tauopathies. Solving the riddle of gait disorders in parkinsonism may require a unifying view that transcends clinicopathological labels.

## Data Availability

The data that support the findings of this study are available from the corresponding author, upon reasonable request.

## Acknowledgments

We acknowledge all our patients and families for actively participating in the study.

## Data Availability

- The authors confirm that the data supporting the findings of this study are available within the article [and/or] its supplementary material.
- The data that support the findings of this study are available from the corresponding author, upon reasonable request.

## Appendix

The project is an initiative of Parkinson Research Alliance of India (PRAI) and Medgenome Labs Collaboration.

